# Tinnitus risk factors and its evolution over time: a cohort study

**DOI:** 10.1101/2024.08.02.24311367

**Authors:** L. Hobeika, M. Fillingim, C. Tanguay-Sabourin, M. Roy, A. Londero, S. Samson, E. Vachon-Presseau

## Abstract

**Background:** Subjective tinnitus is an auditory percept unrelated to an external sound source. The lack of curative treatments and limited understanding of its risk factors complicate the prevention and management of this distressing symptom. This study seeks to identify socio-demographic, psychological, and health-related risk factors predicting tinnitus presence (how often individuals perceive tinnitus) and severity separately, and their evolution over time.

**Methods:** Using the UK Biobank dataset which encompasses data on the socio-demographic, physical, mental and hearing health from more than 170,000 participants, we trained two distinct machine learning models to identify risk scores predicting tinnitus presence and severity separately. These models were used to predict tinnitus over time and were replicated in 463 individuals from the Tinnitus Research Initiative database.

**Finding:** Machine learning based approach identified hearing health as a primary risk factor for the presence and severity of tinnitus, while mood, neuroticism, hearing health, and sleep only predicted tinnitus severity. Only the severity model accurately predicted the evolution over nine years, with a large effect size for individuals developing severe tinnitus (Cohen’s *d* = 1.10, AUC-ROC = 0.70). To facilitate its clinical applications, we simplified the severity model and validated a five-item questionnaire to detect individuals at risk of developing severe tinnitus.

**Interpretation:** This study is the first to clearly identify risk factors predicting tinnitus presence and severity separately. Hearing health emerges as a major predictor of tinnitus presence, while mental health plays a crucial role in its severity. The successful prediction of the evolution of tinnitus severity over nine years based on socio-emotional, hearing and sleep factors suggests that modifying these factors could mitigate the impact of tinnitus. The newly developed questionnaire represents a significant advancement in identifying individuals at risk of severe tinnitus, for which early supportive care would be crucial.

**Funding:** Horizon Europe Marie Slodowska-Curie Actions, the Fondation des gueules cassées, the Fondation pour l’Audition, the Louise and Alan Edwards Foundation, the Canadian Institutes Health Research, the Institut TransMedTech and the Canada First Research Excellence Fund.

## Introduction

Subjective tinnitus is an auditory symptom characterized by the perception of sound without any external acoustic stimulus.^1^ This symptom is common, with a prevalence of 14% in the general population^2,3^, but its severity is highly variable.^4,5^ Tinnitus is not bothersome for most individuals, but it is highly distressing for others who experience sleep disorders, socio-emotional disturbances (i.e. anxiety, depression), and cognitive difficulties.^6^ Because there is no cure for tinnitus but only palliative interventions aiming at reducing associated distress^1,7^, improving tinnitus prevention and clinical management by identifying the key associated risk factors is crucial.

Analogous to phantom limb pain, which is the discomfort or pain felt at the site of an amputated limb, subjective tinnitus is considered a phantom sound following hearing loss. Auditory peripheral damage results in sensory deafferentation within a specific auditory range corresponding to the tinnitus pitch^8^ The auditory loss can be triggered by a variety of causes, including presbycusis, over-exposure to noise, or auditory trauma amongst others. However, this pathophysiological explanation remains unsatisfactory as not everyone with hearing loss experiences tinnitus, and also fails to explain the distress associated with tinnitus. Observed discrepancies in tinnitus experiences may be due variations in socio-demographic, psychological, hearing, or physical health.^9–11^ Moreover, emotional and sleep disorders, often seen as consequences of tinnitus, may also be risk factors for its apparition or severity. In this case, psychosocial factors may instead contribute to shape how tinnitus is experienced by the patient. A longitudinal examination of the risk factors predicting the different facets of tinnitus is currently lacking.

To identify factors predicting the onset and evolution of tinnitus over time, we applied machine learning algorithms to data from the UK Biobank dataset. This extensive biomedical database contains longitudinal detailed information on lifestyle, socio-economic background, hearing, physical and mental health from over 170 000 individuals. As tinnitus presence is not necessarily associated with severity^4^, we analyzed data with two distinct models to predict i) tinnitus presence and ii) its associated severity level, using a pipeline recently developed to study chronic pain. ^12^ We also used the Tinnitus Research Initiative (TRI) dataset to validate our models.^12^ This data driven approach allowed us to tackle three main objectives: 1) determine the risk factors of tinnitus presence and severity, 2) Test if these factors predict their evolution over time 3) Develop a clinical questionnaire to improve prevention and management.

## Methods

### UK Biobank dataset

The UKB dataset is a comprehensive and forward-looking collection of data. More details can be found at https://www.ukbiobank.ac.uk/media/gnkeyh2q/study-rationale.pdf.

#### Participants

Participants aged between 40 and 69 years old, who consented to participate in the study, underwent evaluation at one of the 22 assessment centers in UK. A subset of the participants was subsequently invited for follow-up visits. We used data from the initial assessment (V1) and one of the follow-ups (V2, named Imaging visit in the UKB), with a median time of 9 years between the two visits.

#### Tinnitus phenotypes in the UK Biobank

Tinnitus presence was assessed by: “Do you get or have you had noises (such as ringing or buzzing) in your head or in one or both ears that lasts for more than five minutes at a time?”. Answers were: *(1) Yes. now most or all of the time, (2) Yes. now a lot of the time, (3) Yes. now some of the time, (4) Yes, but not now. but have in the past, (5) No. never, (6) Do not know, (7) Prefer not to answer*. Participants who answered *(4), (6) or (7)* at V1 were excluded. Participants who answered *(6) or (7)* at V2 were excluded. A new category “No, not now” was constituted for V2 to include participants who answered (*4) Yes, but not now. but have in the past*, and *(5) No. never* to include possible recoveries.

All participants who reported experiencing tinnitus were asked: “How much do these noises worry, annoy or upset you when they are at their worst?”. Possible answers were: *(1) Severely, (2) Moderately, (3) Slightly, (4) Not at all, (5) Do not know, (6) Prefer not to answer*. Participants answering *(5) or (6)* at V1 or V2 were excluded from the analysis.

#### Feature selection

We selected 101 features based on their relevance to tinnitus (more details in Table A1). Variables were organized into eleven categories forming four distinct domains, as follows:

##### Hearing health

one category with the items: speech-in-noise hearing test, self-reported deafness, self-reported hearing difficulties with or without noise, medical devices (hearing aid or cochlear implants), and noise exposure.

##### Mood

includes three categories (1) neuroticism, based on 12 neurotic behaviors such as irritability, nervous and guilty feelings; (2) traumas (illness, injury, bereavement or stress in the last 2 years); and (3) mood (reported frequency of certain moods in the past 2 weeks and visits to a GP or psychiatrist for nerves, anxiety, tension or depression).

##### Physical health

includes four categories (1) physical activity based on the Metabolic Equivalent Task (MET) scores computed using the International Physical Activity questionnaire (IPAQ)^13^; (2) sleep; (3) substance use (smoking and alcohol); and (4) anthropometric measures such as BMI, fractures and blood pressure.

##### Sociodemographic

includes three categories (1) socioeconomic status, such as education, income and employment; (2) occupational measures, such as social entourage and manual or physical job; and (3) demographics such as age, sex and ethnicity.

#### Missing data

Since hearing evaluation was added to V1 a few years after the initial data collection began, we included in this study the 172,451 out of 493,211 participants who had complete hearing data. Individuals with more than 20% of missing data for the 101 predictors were excluded. For the others, missing data were replaced by the feature median. Features were standardized across participants by centering the mean to zero and scaling the variance to one.

### Data analyses in the UK Biobank

#### Developing the predictive models of tinnitus presence and tinnitus severity

We used the NIPALS regression algorithm (implemented using scikit-learn.org/) on the 101 features to create risk scores predicting separately (1) Tinnitus presence (Figure 1.A) and (2) Tinnitus associated severity (Figure 2.A). NIPALS identifies latent patterns that maximize the covariance between two matrices (details in appendix A1). To this end, the UK Biobank dataset at V1 was divided into a training set (*n* = 147,133 for the presence model, *n* = 43,906 for the severity model) for discovery and a testing set composed of out-of-sample participants for whom longitudinal data were available (*n* = 20,850 for the presence model, *n* = 4,291 for the severity model). The algorithms were trained using tenfold cross-validation to estimate the models. The trained models were then applied to the participants of the testing set. The two models’ output provided a single prediction for tinnitus presence and its associated severity separately, for each participant. These outputs are referred to as the risk score for tinnitus presence and the risk score for tinnitus severity.

**Figure 1:**
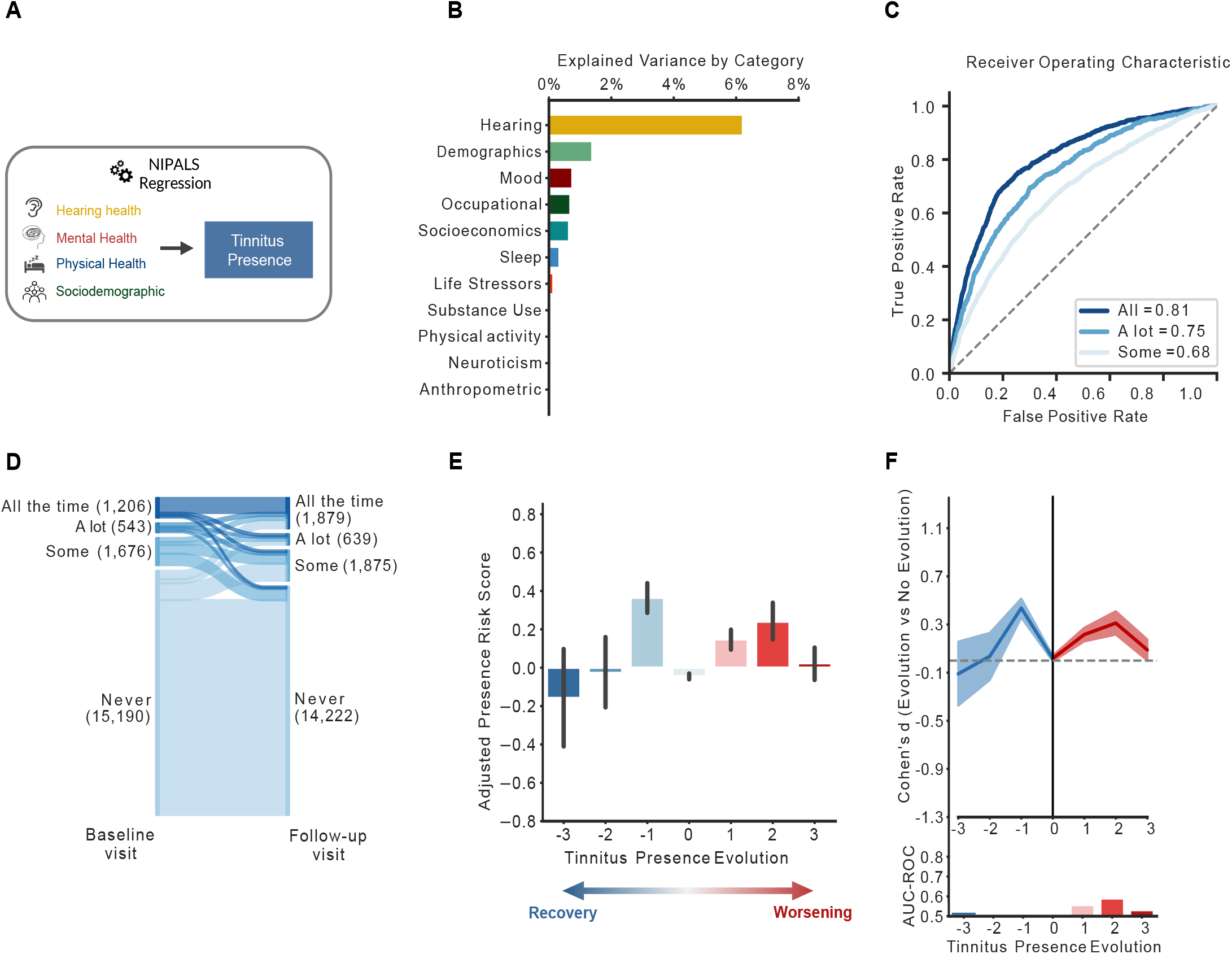
Tinnitus presence model. **A**. We used the NIPALS machine learning algorithm to predict the presence of tinnitus based on 101 features, representing four categories: hearing health, mental health, physical health and sociodemographic. **B**. The explained variance for all subcategories is depicted. Only hearing and demographic factors explained more than 1% of the variance each. **C**. The model had good to excellent performances to classify participants based on how often they experienced tinnitus (some of the time, a lot of the time, all the time), as shown by the AUC-ROC curves. **D**. This panel depicts the evolution of tinnitus presence between the baseline visit (left side) and the follow-up visit (right side), spaced by nine years. **E and F**. Those panels showed the evolution of the adjusted risk scores (**E**), and the performances (Cohen’s d and AUC-ROC) of the model (**F**) in function of the evolution of tinnitus presence over time. The evolution is rated between −3 and 3, with −3 representing the evolution from tinnitus present all the time at baseline to the absence of tinnitus in the follow-up visit, and +3 the opposite evolution. Based on those figures, we concluded that the model could not predict the evolution of tinnitus presence over time.

**Figure 2:**
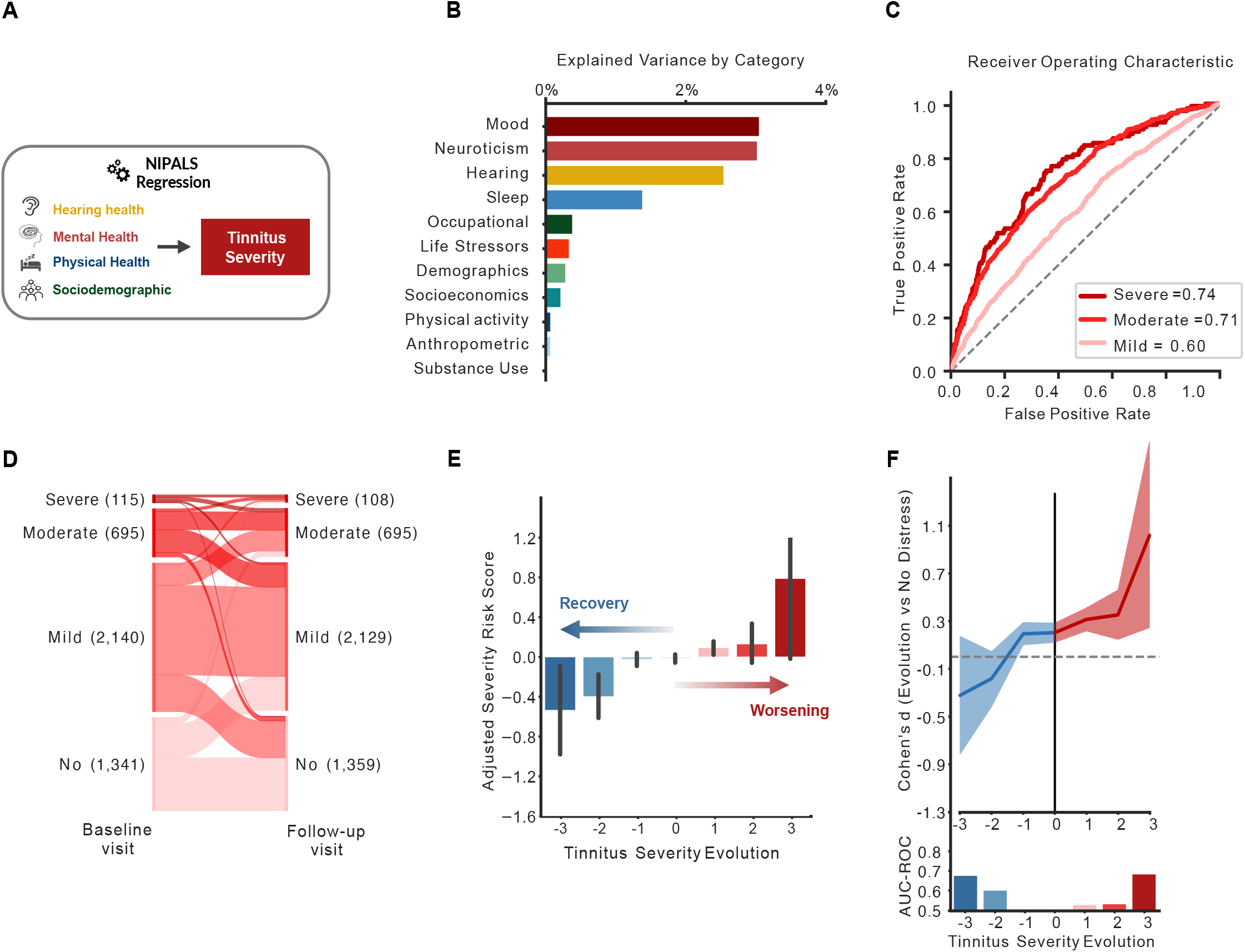
Tinnitus severity model. **A**. We used the NIPALS machine learning algorithm to predict the severity of tinnitus based on 101 features, representing four categories: hearing health, mental health, physical health and sociodemographic. **B**. The explained variance for all subcategories is depicted. The mood, neuroticism, hearing and sleep explained more than 1% of the variance each. **C**. The model had good performances to classify participants experiencing a moderate to severe tinnitus associated distress, as shown by the AUC-ROC curves. **D**. This panel depicts the evolution of tinnitus severity between the baseline visit (left side) and the follow-up visit (right side), spaced by nine years. **E and F**. Those panels showed the evolution of the adjusted risk scores (**E**), and the performances (Cohen’s d and AUC-ROC) of the model (**F**) in function of the evolution of tinnitus severity over time. The evolution is rated between −3 and 3, with −3 representing the evolution from a severe distress to no distress, and +3 the opposite evolution. Based on those figures, we concluded that the model had good performances in predicting the evolution of tinnitus over time for the −3 and +3 categories.

#### Tinnitus evolution over time

The prognostic values of the tinnitus presence and severity risk scores to predict the recovery, persistence or worsening of tinnitus were assessed using the testing datasets. We created adjusted risk scores, which were orthogonal to the baseline tinnitus level, to interpret if interindividual deviations predicted the evolution over time (details in Appendix A2)

#### Tinnitus severity evolution simplified risk score

A simplified model of the tinnitus severity risk score was derived from the full risk model using the training dataset. Non-modifiable factors (sex, age and ethnicity), quantitative measures (hand grip strength, hearing test) and composite scores (total neuroticism score, number of life stressors) were excluded from this simplified model to include only modifiable, easily collectable declarative items. We trained a linear forward feature selection algorithm, implemented using scikit-learn, to select the core features that captured the highest explained variance. Features are iteratively added to the model for a prespecified combination of features in the 101 features pool until there is no improvement in the model’s performance. We used the elbow method to determine the number of features providing the best trade-off between sparsity and variance explained.

#### Simplified risk score validation: TRI Database

The TRI dataset is composed of 4,246 individuals with tinnitus who answered questionnaires about their tinnitus, their physical, hearing and mental health while visiting a German tinnitus clinic between 2010 and 2023.^14^ A subset of 463 individuals, aged between 25 and 86 years old, came back for at least one follow-up visit, with a median time of 4 years between the two visits. We used the Tinnitus Handicap Inventory^15^ (THI) scores and categorization to determine participants tinnitus severity (No, Mild, Moderate, Severe, Catastrophic). An equivalent to the Tinnitus severity evolution simplified risk score was constructed using questions of the TRI database; to determine both the classification and longitudinal validity of the score (details in appendix A3).

#### Statistical analysis

The models fit were assessed using the explained variance (*R*^2^). The risk scores of individuals with different levels of tinnitus presence or severity were compared to the score of tinnitus free participants for the presence, and distress-free tinnitus participants for the severity, using Cohen’s *d* effect sizes and AUC-ROCs. We used bootstrap resampling with 10,000 iterations to indicate the estimated error in the Cohen’s *d* effect sizes. Analyses were performed using Python v.3.11.5 with Spyder 5.4.3, including Numpy (v.1.24.3), Pandas (v.2.0.3), Sklearn (v.1.3.0), Seaborn (v.0.12.2), Matplotlib (v.3.7.2), Pingouin (v.0.5.3) and Nltools (v.0.5.0).

#### Ethical approval

The UKB was approved by the Research Ethics Committee (no. 11/NW/0382). Ethical approval for the collection of the TRI database was obtained from the Ethics Committee of the University of Regensburg (protocol number 08/046). All participants gave written, informed consent.

## Results

### Descriptive

Table 1 shows the prevalence of tinnitus presence and the severity categorization among participants with tinnitus. The percentage of participants with tinnitus was 20.0%, with 20.2% of them experiencing a moderate or severe distress. Tinnitus presence was more prevalent in men, but more distressing for women.

**Table 1:** Baseline characteristics at the baseline visit. Data are percentage, except for the Age and Speech reception Threshold for which the mean and (standard deviation) are given.

|  | Tinnitus Presence (N = 167,983) |  |  |  | Tinnitus Severity (N = 48,197) |  |  |  |
| --- | --- | --- | --- | --- | --- | --- | --- | --- |
|  | Never had | Some of the time | A lot of the time | All the time | No distress | Mild distress | Moderate distress | Severe distress |
| **Prevalence** | 80.0 % | 10.1 % | 2.7 % | 7.2 % | 32% | 47.8 % | 16.6 % | 3.6 % |
| **Demographics** |  |  |  |  |  |  |  |  |
| **Sex (% M/F)** | 43.8 / 56.2 | 48.0 / 52.0 | 53.7 / 46.3 | 59.3 / 40.7 | 55.3 / 44.7 | 48.2 / 51.8 | 46.3 / 53.7 | 43.1 / 56.9 |
| **Age** | 56.4 (8.2) | 57.9 (7.8) | 58.9 (7.3) | 59.6 (7.2) | 57.0 (8.3) | 57.9 (7.8) | 58.1 (7.8) | 57.3 (7.8) |
| **Hearing Health** |  |  |  |  |  |  |  |  |
| **Self-reported hearing difficulties** | 19.5 % | 45.1 % | 59.2 % | 70.5 % | 37.2 % | 47.7 % | 62.6 % | 66.9 % |
| **Self-reported speech-in-noise hearing difficulties** | 29.9 % | 52.8 % | 61.7 % | 67.5 % | 44.5 % | 55.0 % | 66.3 % | 69.3 % |
| **Speech Reception Threshold in dB** | -6.6 (1.9) | -6.3 (2.4) | -6.3 (2.5) | -6.0 (2.5) | -6.5 (2.2) | - 6.3 (2.3) | - 6.0 (2.5) | - 5.6 (3.2) |
| **Hearing aid users** | 1.7 % | 5.5 % | 7.0 % | 12.1 % | 4.5 % | 5.4 % | 9.7 % | 14.6 % |
| **Cochlear implants** | .060 % | .32 % | .17 % | .31 % | .17 % | .19 % | .34 % | .82 % |

### Tinnitus presence risk score: classification at baseline and evolution in time

#### Risk score calculation

The model explained a total of 11% of the variance of the tinnitus presence, with the most explained variance coming from hearing health (6.2%) followed by demographic (1.4%), whereas other categories explained the least variance (<1% each) (Figure 1.B). The weights of each feature in the model are presented in the Appendix (Figure A1). The risk score for tinnitus presence showed good to excellent performance for classifying participants with tinnitus from tinnitus-free participants, as shown by their diagnostic capacities (AUC 0.68–0.81, Figure. 1C) and effect sizes (Cohen’s *d* = 0.70, 1.05, 1.36 for the levels some, a lot and all the time respectively). We confirmed the validity of the risk score independently for different ethnicities (ROC-AUC > 0.71 for all the time vs no tinnitus, for Asian, black and white ethnicities, see Appendix A4).

#### Recovery and worsening over time: 9-year prognosis

Participants evolution of tinnitus presence at the two visits are displayed in Figure.1D. The adjusted presence risk score did not predict the evolution of tinnitus at the follow up visit, as evidenced by the Cohen’s d (all < .50) and AUC-ROC levels (AUC < 0.60, Figure 1.E and F). As the presence risk score did not predict the evolution, we trained a new model specifically to predict the evolution of tinnitus presence over time, using the 101 features. This new model was also unsuccessful at predicting the evolution of tinnitus presence over time (AUC-ROC ≤ .60 for every evolution level). This suggests that the evolution of tinnitus presence can hardly be predicted from general health, sociodemographic or environmental factors.

### Tinnitus severity risk score: classification at baseline and evolution in time

#### Risk score calculation

The model explained a total of 7% of the variance of the tinnitus severity, with the most explained variance coming from mood (3.0%), neuroticism (3.0%), hearing health (2.5%) and sleep (1.4%), whereas other categories explained the least variance (<1% each). The weights of each feature in the model are presented in Appendix (Figure A2). The risk score for tinnitus severity showed moderate to excellent performance for classifying participants with distressing tinnitus from distress-free participants, as shown by diagnostic capacities (AUC 0.60–0.74, Figure. 2C) and their effect sizes (Cohen’s *d* = 0.35, 79, 0.90 for the levels mild, moderate and severe respectively). We confirmed the validity of the risk score independently for different ethnicities (ROC-AUC > .71 for severe tinnitus vs no distress, for Asian, black and white ethnicities, see Appendix A4).

#### Recovery and worsening over time: 9-year prognosis

The stability and individual changes in tinnitus severity between the two visits are displayed in Figure.2D. The adjusted severity risk score predicted the evolution of tinnitus at the follow up visit, as seen in the evolution plot (Figure 2.E), the Cohen’s d and AUC-ROC levels (AUC = 0.68 for an evolution from no distress to severely distressing, and AUC = 0.68 for an evolution from severely distressing to no distress Figure 2.F).

#### Evolution of tinnitus severity over time: a clinical questionnaire

Last, we aimed to simplify our model and reduce the number of features by extracting those with the higher predicting value. This simplified model is a reduced risk score for tinnitus severity calculated by simply summing the binarized answers to five items measuring hearing health, sleep, neuroticism and mood, selected with a linear forward feature selection algorithm. The questionnaire gathering those five items is called POST (Prediction Of the Severity of Tinnitus) (Figure. 3A). The simplified risk score achieved good performance in predicting severity (Figure 3, B). It also had an average to good performance to predict tinnitus evolution in the longitudinal dataset (Figure 3, C,D), especially in the prediction of individuals who will evolve from a non-distressing tinnitus to a severely distressing one (Cohen’s *d =* 1.6, ROC = 0.74). This represented a good trade-off between the smallest number of features and the highest AUC-ROC. Based on the odd ratios of experiencing no, mild, moderate or severe at the follow-up visit depending on the Reduced risk score at baseline, we concluded that scores of 0 and 1 are associated with a low risk, 2 and 3 are associated with a moderate risk and 4 and 5 are associated with high risk of experiencing moderate of severe distress over time (Appendix A5).

**Figure 3:**
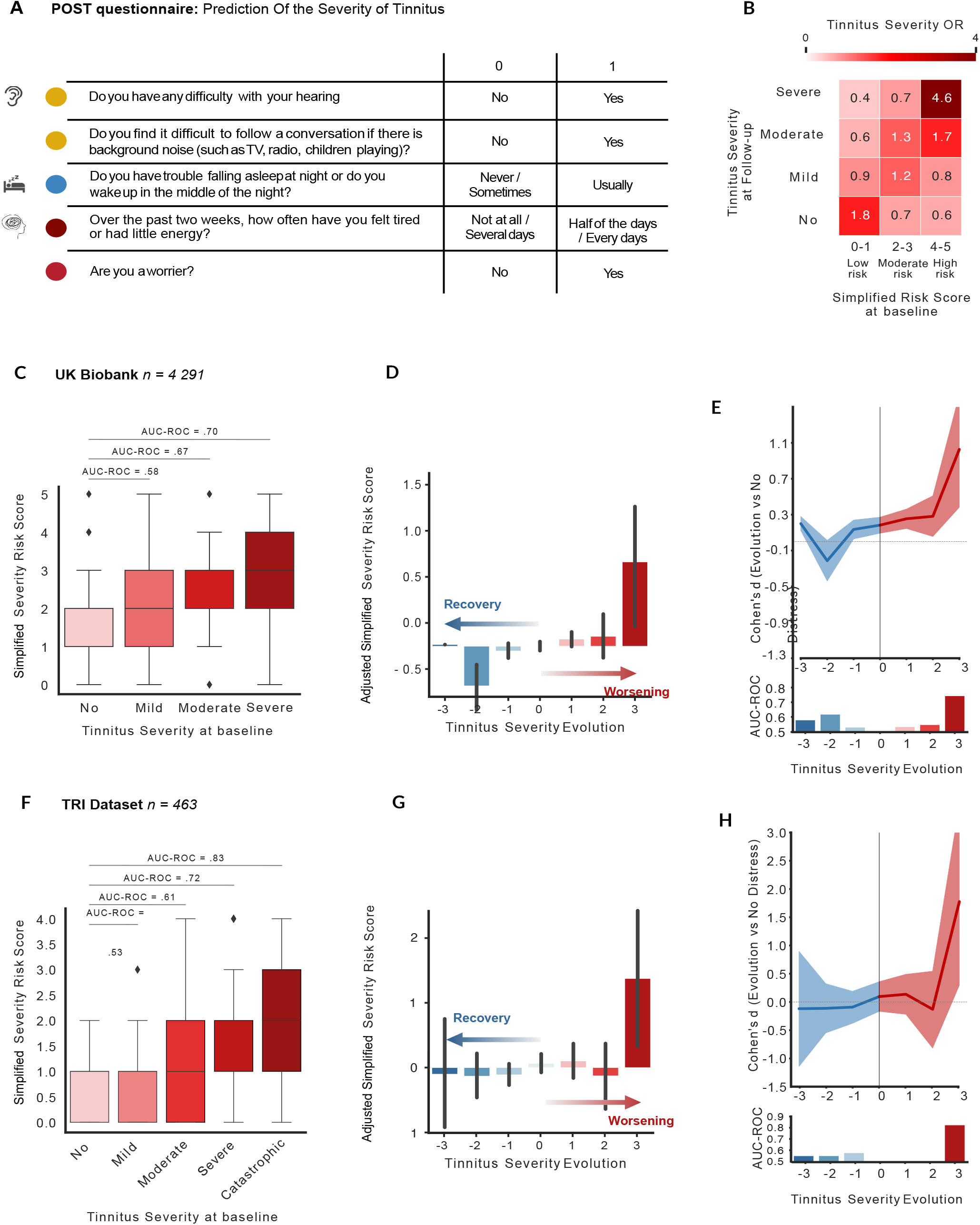
Validation of the POST questionnaire. Based on the severity model, we extracted five items explaining the most variance to create a short questionnaire, with binarized answers to estimate the risk of developing moderate or severe tinnitus over time. The simplified risk score is constituted of two items on hearing health, one on sleep disorders and two on mental health. **B**. This figure depicts the odd ratios of experiencing no, mild, moderate or severe distress associated with tinnitus at a follow-up study based on the risk score at Baseline. Based on those odd ratios, we conclude that a risk scores of 0 and 1 is associated with a low risk of developing moderate or severe distress over time, risk scores of 2 and 3 are associated with a moderate risk, while risk scores of 4 and 5 are associated with a high risk. **C to G**. We tested the validity of this simplified risk score on data of the UK Biobank (**B** to **D**) and of the TRI (**E** to **G**). Figures **B** and **E** evidenced that the simplified risk score had moderate to excellent performances in classifying tinnitus severity at baseline. Figures **C, D, F** and **G** panels showed the evolution of the adjusted risk scores, and the performances (Cohen’s d and AUC-ROC) of the model (**F**) in function of the evolution of tinnitus severity over time. The simplified model had excellent performances to detect individuals at risk of evolving to a severe or catastrophic distress.

We validated our simplified risk score on 463 patients experiencing tinnitus of the TRI dataset. Even though the Severity was evaluated with 5 categories, the evolution was rated between −3 and 3, and not −4 and 4 as there were no individuals who evolved from no distress to an invalidating tinnitus or reversely. Here, the simplified risk score associated with tinnitus severity achieved good performance (Fig 3.F), concordant with what was initially observed in the UKB. POST also had an average to good performance to predict tinnitus evolution in the longitudinal dataset (Figure 3.F.G) especially in the prediction of individuals who will evolve from a non-distressing tinnitus to a severely distressing one (Cohen’s *d =* 1.8, ROC = 0.83).

## Discussion

The objective of this study was to identify the risk factors of tinnitus presence and severity, as well as their evolution over time. The results revealed a dissociation between the features predicting tinnitus presence and tinnitus severity. While hearing health emerged as a common key predictor of presence and severity, mood, neuroticism and sleep only predicted its severity. Interestingly, while the presence model did not predict the evolution of tinnitus over time, the severity model provided an estimation of its progression over nine years, with a large effect size for individuals who develop severe tinnitus. A simplified version of the risk score for tinnitus severity was derived from five questions with binarized outcomes and validated in two independent cohorts with the aim to detect individuals at risk of developing severe tinnitus over time.

In the UK Biobank, tinnitus prevalence was 17.7%, with a moderate to severe severity for 20.3% of them, which is in line with common prevalence observed for this age range (40-70).^2,3^ Tinnitus was more prevalent for men than women, while tinnitus severity was higher in women. Looking at the relationship between tinnitus and all measures of hearing health, we observed increasing deficits with increasing tinnitus presence and with increasing tinnitus severity. Overall, our results indicate that, even if the UK Biobank has potential biases and limitations, such as healthy volunteer selection bias^16^, lack of ethnic diversity (91% of our sample are of white descents), the prevalence of tinnitus is aligned with the literature.^2,17–19^

Various factors have been associated with tinnitus presence or severity, such as mental health, education level, chronotype, physical exercise or alcohol consumption^20–22^, but with low levels of evidence.^10^ These often interrelated factors are usually studied in isolation. To overcome this limitation, we included a large variety of possible risk factors in the same multivariate model, covering socio-demographics, hearing health, mental health and physical health factors, merging them into categories to create a global picture of tinnitus pathophysiology. First, we showed that the major predictor of tinnitus presence is hearing health, and in particular self-reported hearing difficulties. It confirms the large literature pointing toward hearing deficits as the main risk factor for tinnitus apparition.^1^ The second identified risk factor is age, which is likely mediated by presbycusis. Emotional factors have also been identified as potential risk factors in the literature^2^, attributed to so called stress-induced tinnitus.^23^ Our results suggest that emotional factors explain only a small part of the variance, showing limited predictive capacities. Physical health factors, like anthropometrics, physical activity and substance use had no predictive value of tinnitus presence. Overall, our results indicate that tinnitus presence is mainly predicted by hearing health. They do not explain the fact that not all individuals with hearing loss develop tinnitus. In order to clarify this observation, it is essential to look into biological factors such as genetics^24^ and cerebral functioning^25^ which extend beyond the scope of this study.

Our results show that tinnitus severity is predicted mainly by: mood (anxiety, depression), neuroticism (i.e. personality trait characterized by a tendency to respond with negative emotions to threat, frustration or loss^26^), sleep and life stressors. This result is in line with the literature that has extensively associated severe tinnitus with stress, depression, personality traits and sleep disorders.^1,11^ On the other hand, the association between severity and hearing health has been reported more rarely in the literature.^27,28^ This could reflect differences in tinnitus loudness or masking level. Other potential risk factors identified in the literature, like physical activity or substance use, show very small predictive value. We hypothesized that these effects are mediated by other socio-emotional factors.

After identifying risk factors, our main challenge was to understand the factors mediating tinnitus evolution over time. The risk score for tinnitus presence was unable to predict the evolution in the levels of tinnitus presence over time, which was expected, as the risk score primarily reflects hearing deficits may either not have yet occurred or are largely irreversible. This suggests that risk factors for the evolution of tinnitus presence may be the result of pathophysiological mechanisms in the auditory periphery or the central nervous system rather than from psychosocial factors. On the contrary, we showed that mood, personality traits, sleep, and hearing dysfunction were the strongest predictors of the evolution of tinnitus severity over time. This aligns with previous studies associating tinnitus severity with mood and sleep disorders, but for the first time, we demonstrate that these factors actually predispose tinnitus severity evolution, evidencing a key role of those factors in the prediction of its progression. This implies that these factors, by the means of sound or psychological oriented therapies, may not stop tinnitus perception *per se* but may instead alleviate how they are experienced. Those results are in coherence with the clinical approach of decreasing the severity instead of stopping the percept.

To improve clinical utility, we finally created a 5-items questionnaire that predicts tinnitus severity over time. This represents an easy-to-use prognostic tool for identifying patients who are unlikely to habituate to tinnitus. This questionnaire gives good results identifying out of sample participants at risk of developing severe tinnitus. These results were validated in an independent clinical dataset (TRI database, 462 participants), ensuring the generalization of our results. This questionnaire can be a key tool to improve tinnitus clinical management at an earlier stage, by concentrating the limited resources, few healthcare professionals trained in severe tinnitus management^29^, for patients at risk of developing severe tinnitus, while avoiding unnecessary or oversized interventions for those more likely to habituate.^8^

Overall, our study clearly distinguishes between the presence and severity of tinnitus, which is in line with the definition proposed by an international multidisciplinary consortium who distinguish between tinnitus (the perception of a sound without external source), and tinnitus disorder (tinnitus with distress).^4^ Additionally, we show that only tinnitus severity can be predicted, pinpointing differences in risk factors associated with each condition. Tinnitus, often associated with hearing health, highlights the necessity for raising public awareness about the irreversible consequences of peripheral auditory damage induced by noise or ototoxic drugs. Conversely, tinnitus disorder is influenced by psychosocial factors, underscoring the significance of interventions targeting these factors.

## Data Availability

The dataset from UK Biobank analyzed in the study is available via application to the Access Management System athttps://www.ukbiobank.ac.uk . The TRI dataset is accessible upon requests, see https://tinnitusresearch.net/index.php/for-researchers/tinnitus-database .

## Acknowledgments

LH was funded by the Horizon Europe Framework Programme (HORIZON) under the Marie Sklodowska-Curie Postdoctoral Fellowship (grant No. 101146406) and the Fondation des Gueules Cassées. LH and SS were supported by the Fondation pour l’Audition (FPA RD-2019-10). MF was funded by the Louise and Alan Edwards Foundation. CTS was funded by the Canadian Institutes of Health Research, the Institut TransMedTech and the Canada First Research Excellence Fund.

## Contributors

All authors conceived and designed the study. LH, MF and CTS created the methodology. LH and MF performed the statistical analyses. LH drafted the manuscript. All authors were involved in the interpretation of data, and critical revision of the manuscript. All authors had full access to all of the data.

## Data sharing

The dataset from UK Biobank analyzed in the study is available via application to the Access Management System at https://www.ukbiobank.ac.uk. The TRI dataset is accessible upon requests, see https://tinnitusresearch.net/index.php/for-researchers/tinnitus-database.

## Declaration of generative AI and AI-assisted technologies in the writing process

During the preparation of this work, the authors used ChatGPT 3.5 / OpenAI in order to improve the flow and readability of the writing. After using this tool, the authors reviewed and edited the content as needed and takes full responsibility for the content of the publication.

