## Supplementary materials for "Tinnitus risk factors and its evolution over time: a cohort study"

### A1. Developing the predictive models predicting tinnitus presence and severity

The nonlinear iterative partial least square (NIPALS) [27] method was trained within the discovery dataset to identify latent scores (**E** and **Z**) and loadings (**P** and **H**) that maximize the covariance between matrix of standardized psychosocial features ($X_i$) (size (147385, 101) for tinnitus presence, and (43983, 101) for tinnitus severity) and a vector of self-reported tinnitus presence or severity (**Y**) (size (147385, 1) for tinnitus presence, and (43983, 1) for tinnitus severity):

1. Compute singular vectors **μ**, **ν** (weights) of covariance matrix $C = X^T\mathbf{Y}$
2. Obtain the latent scores **E** and **Z** by projecting $X$ and **Y** onto singular vectors **μ** and **ν**
3. Compute loadings **P** and **H** by iteratively regressing $X$ onto **E** (power iteration)
4. 4. Deflate $X$ and **Y** using $X + 1 = X - \mathbf{E}\mathbf{P}^T$ and $\mathbf{Y} + 1 = Y - \mathbf{Z}\mathbf{H}^T$, respectively
5. 5. Fit training (discovery) data $X$ using the projection matrix **P** to obtain latent space $\bar{x}$ so that $\bar{x} = X\mathbf{P}$
6. Use the latent space to predict left-out data $\mathbf{Y_V}$ (size (20853,1) for tinnitus presence, (4291,1) for tinnitus severity) using the coefficient matrix $\beta \in R^{d \times t}$ $\beta$ such that $\mathbf{Y_V} = \mathbf{X_V}\beta$, where $X_V$ denotes the matrix of psychosocial features in the validation set ($X_V$ size (20853,101) for tinnitus presence, (4291,101) for tinnitus severity)

Further information on the implementation can be found at https://scikit-learn.org/stable/modules/cross_decomposition. html#cross-decomposition.

### A2. Adjustment of the risk scores for the longitudinal evaluations

To examine the prognostic value of the presence risk score, we regressed out the squared values of the tinnitus presence level (0: No, 1: Some of the time, 2: A lot of the time, 3: All of the time) at baseline from presence the risk score calculated at baseline. Making the score orthogonal to the Tinnitus presence level at baseline allowed us to interpret interindividual deviations in this adjusted score as risk of recovery or worsening of tinnitus presence at the follow-up visit. Similar analysis was performed on the severity risk score with the levels 0: No, 1: Mild, 2: Moderate and 3: Severe). The process is depicted in the following figure.

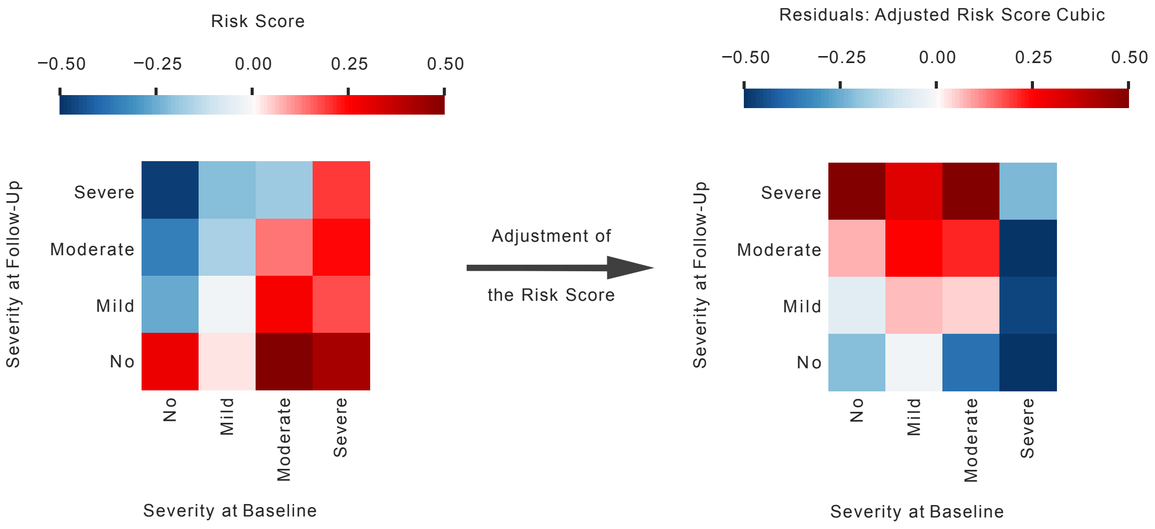

### A3. Reduced risk score replication: TRI Database

The reduced severity risk score was derived using five binarized items:

1. **Do you think you have a hearing problem?** 0: No / 1: Yes
2. **Do you wear hearing aids** 0: No / 1: Yes
3. **How much of the time have you had trouble sleeping at night?** 0: At no time, 0: Some of the time, 0: Slightly less than half the time / 1 = Slightly more than half the time, 1: Most of the time, 1: All the time
4. **How much of the time have you felt lacking in energy and strength?** 0: At no time, 0: Some of the time, 0: Slightly less than half the time / 1 = Slightly more than half the time, 1: Most of the time, 1: All the time
5. **How much of the time have you had a bad conscience or feelings of guilt?** 0: At no time, 0: Some of the time, 0: Slightly less than half the time / 1 = Slightly more than half the time, 1: Most of the time, 1: All the time

The TRI database did not contain information on speech-in-noise hearing difficulties, which could be a proper equivalent of the original item "Do you find it difficult to follow a conversation if there is background noise (such as TV, radio, children playing)?". We replaced it by the hearing health item having the larger weight in the severity model, which is "Do you wear hearing aids?".

### A4: Validation of the presence and severity risk scores over ethnicities

To ensure the validity of the presence and severity models on different ethnicities, we tested the performances of the models separately on groups of Asian, Black and White individuals. We tested participants which were not included in the rest of the analysis, using data of participants who did not had a full hearing evaluation at baseline visit but had one in the follow-up visit. We tested 466 Asian, 203 Black and 41,969 White individuals with the presence model, and 81 Asian, 38 Black and 10,115 White individuals with the severity model. The presence model **(A)** had good to excellent performances to classify individuals perceiving tinnitus a lot or all the time (except for the Asian "a lot of the time" group). The severity model **(B)** had good to excellent performances to classify individuals with moderate or severe distress (except for the Asian "moderate" group). Overall, the models had good performances for the extreme levels (all the time, or severe distress) for all ethnicities. The performances for the intermediate levels (A lot of the time, moderate distress) were good for Black and White individuals, but low for Asian. Those results should be replicated on larger groups of individuals.

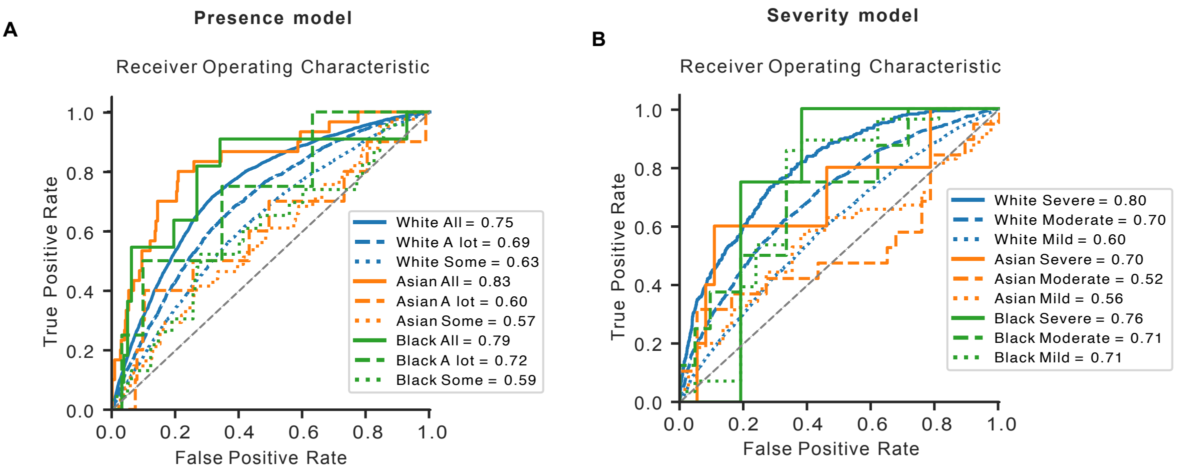

**A5. Examining the severity at follow-up depending on the reduced risk score at baseline**

This figure depicts the odd ratios of experiencing a severe, moderate, mild or no distress associated with tinnitus at the follow-up visit, depending on the Reduced risk score at baseline. Based on those odd ratios, we can conclude that 0 and 1 risk scores are associated with a low risk of experiencing moderate or severe tinnitus over time. Scores 2 and 3 are associated with a moderate risk, and score 4 and 5 are associated with a large risk of experiencing moderate or severe tinnitus distress over time.

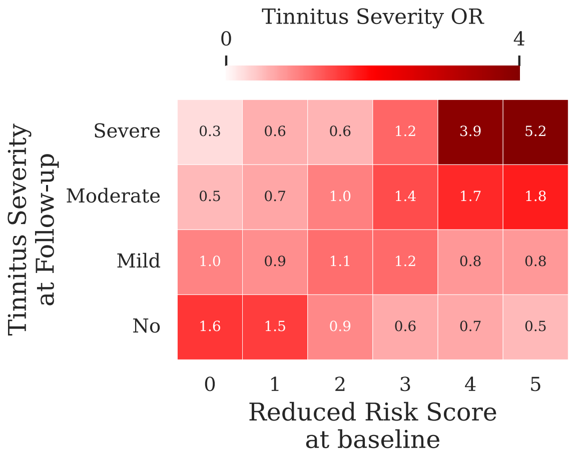

**Table A1:** Variables extracted from the UK Biobank and their associated data field.

| Type of measure | Details | UK Biobank Data Field |
|---|---|---|
| **Tinnitus evaluation** | | |
| Tinnitus | | 4803 |
| Tinnitus severity/nuisance | | 4814 |
| **Hearing** | | |
| Hearing difficulties self-reported | | 2247 |
| Hearing difficulties self-reported with background noise | | 2257 |
| Hearing test right ear | | 20021 |
| Hearing test left ear | | 20019 |
| Hearing aid | | 3393 |
| Cochlear implants | | 4792 |
| Noisy workplace | | 4825 |
| Loud music exposure | | 4836 |
| **Mood** | | |
| Frequency of depressed mood in last 2 weeks | | 2050 |
| Frequency of unenthusiasm/disinterest in last 2 weeks | | 2060 |
| Frequency of tenseness/restlessness in last 2 weeks | | 2070 |
| Frequency of tiredness/lethargy in last 2 weeks | | 2080 |
| Seen doctor (GP) for nerves, anxiety, tension or depression | | 2090 |
| Seen psychiatrist for nerves, anxiety, tension or depression | | 2100 |
| Risk taking | Whether the participant describes themselves as someone who takes risks | 2040 |
| **Neuroticism** | | |
| Neurotic behaviors | Mood swings | 1920 |
| | Miserableness | 1930 |
| | Irritability | 1940 |
| | Sensitivity/hurt feelings | 1950 |
| | Fed-up feelings | 1960 |
| | Nervous feelings | 1970 |
| | Worrier/anxious feelings | 1980 |
| | Tens/highly strung | 1990 |
| | Worry too long after embarrassment | 2000 |
| | Suffer from nerves | 2010 |
| | Loneliness | 2020 |
| | Guilty feeling | 2030 |
| Total neuroticism score | Derived from the 12 neurotic behaviors, as a sum score of the total "yes" answers to these questions | 20127 |
| **Life stressors** | | |
| Serious illness, injury, bereavement in the last 2 years | Serious trauma to self<br>Serious trauma to close relative<br>Death of close relative<br>Death of spouse or partner<br>Martial separation/ divorce<br>Financial difficulties | 6145 |
| Life stressors whithin last 2 years | Derived from the life stressors 6145, as a sum score of the total "yes" answers to these questions | - |
| **Sleep** | | |
| Sleep duration | Hours of sleep in every 24 hours | 1160 |

| | | |
|---|---|---|
| Getting up in the morning | Difficulty getting up | 1170 |
| Sleeplessness/insomnia | Trouble falling asleep at night or waking up in the middle of the night | 1200 |
| Nap during the day | | 1190 |
| Chronotype | Late chronotype (evening person)<br>Early chronotype (morning person) | 1180 |
| Daytime dozing (narcolepsy) | Likely to doze off or fall asleep during the daytime | 1220 |
| **Physical activity** | | |
| Hand grip strength | Average between left- and right-hand grip strength (units of measurement: Kg). | 46 & 47 |
| IPAQ activity group | Low<br>Moderate<br>High | 22032 |
| MET minutes per week for walking | Units of measurement: minutes/week | 22037 |
| MET minutes per week for moderate activity | Units of measurement: minutes/week | 22038 |
| Above moderate/vigorous recommendation | Indicates if the participant met the 2017 UK Physical activity guidelines of 150 minutes of moderate activity per week or 75 minutes of vigorous activity | 22035 |
| **Substance Use** | | |
| Smoking status | Previous smoker | 20116 |
| Current tobacco smoking | Daily smoker<br>Occasional smoker | 1239 |
| Past tobacco smoking | How often the participant smoked tobacco in the past | 1249 |
| Ever smoked | Derived from current and past tobacco smoking status (data fields 1239 & 1249) | 20160 |
| Smokers in household | If anyone smokes in the participant's household | 1259 |
| Hours of exposure to tobacco at home | Hours per week | 1269 |
| Alcohol drinker status | Never drank<br>Previous drinker | 20117 |
| Alcohol intake frequency | | 1558 |
| Alcohol intake vs. 10 years previously | Indicates the change in the participant's alcohol use compared to 10 years ago | 1628 |
| **Anthropometric** | | |
| Body mass index (BMI) | Constructed from height and weight measured during the in-person assessment visit | 21001 |
| Weight change compared with one year ago | Gained weight<br>Lost weight | 2306 |
| Weight | Unit of measurement: kg | 21002 |
| Systolic blood pressure | Units of measurement: mmHg | 4080 |
| Diastolic blood pressure | Units of measurement: mmHg | 4079 |
| Pulse rate | Pulse rate measured during the automated blood pressure readings (units of measurement: bpm). | 102 |
| Fractured/broken bones in last 5 years | | 2463 |
| **Occupational** | | |
| Job involves heavy manual or physical work | | 816 |
| Job involves walking or standing | | 806 |
| Current employment status | Looking after home or family<br>Paid employment/self-employed<br>Retired<br>Unemployed<br>Unable to work or disable | 6142 |
| Education (qualifications) | College or university<br>Advanced level | 6138 |

| | | |
|---|---|---|
| | Ordinary level<br>Certificate secondary education<br>Practical career diploma<br>Other professional qualifications | |
| ***Demographics*** | | |
| Ethic background | White<br>Asian<br>Black<br>Mixed<br>Other | 21000 |
| Sex | Female / Male | 31 |
| Age | Age in years | 21003 |
| **Socioeconomics** | | |
| Number in household | Number of people living together in participant's household including themselves | 709 |
| Relationship with people living in household | Living with partner<br>Living with children<br>Living with siblings<br>Living with parents<br>Living with grandparents<br>Living with grandchildren<br>Living with related relatives<br>Living with unrelated relatives | 6141 |
| Frequency of friends and family visits | | 1031 |
| Able to confide | How often the participant has been able to confide to someone close to them | 2110 |
| Number of vehicles in household | | 728 |
| Average total household income | | 738 |

**Figure A1:** Features weights of the Presence Risk Score model

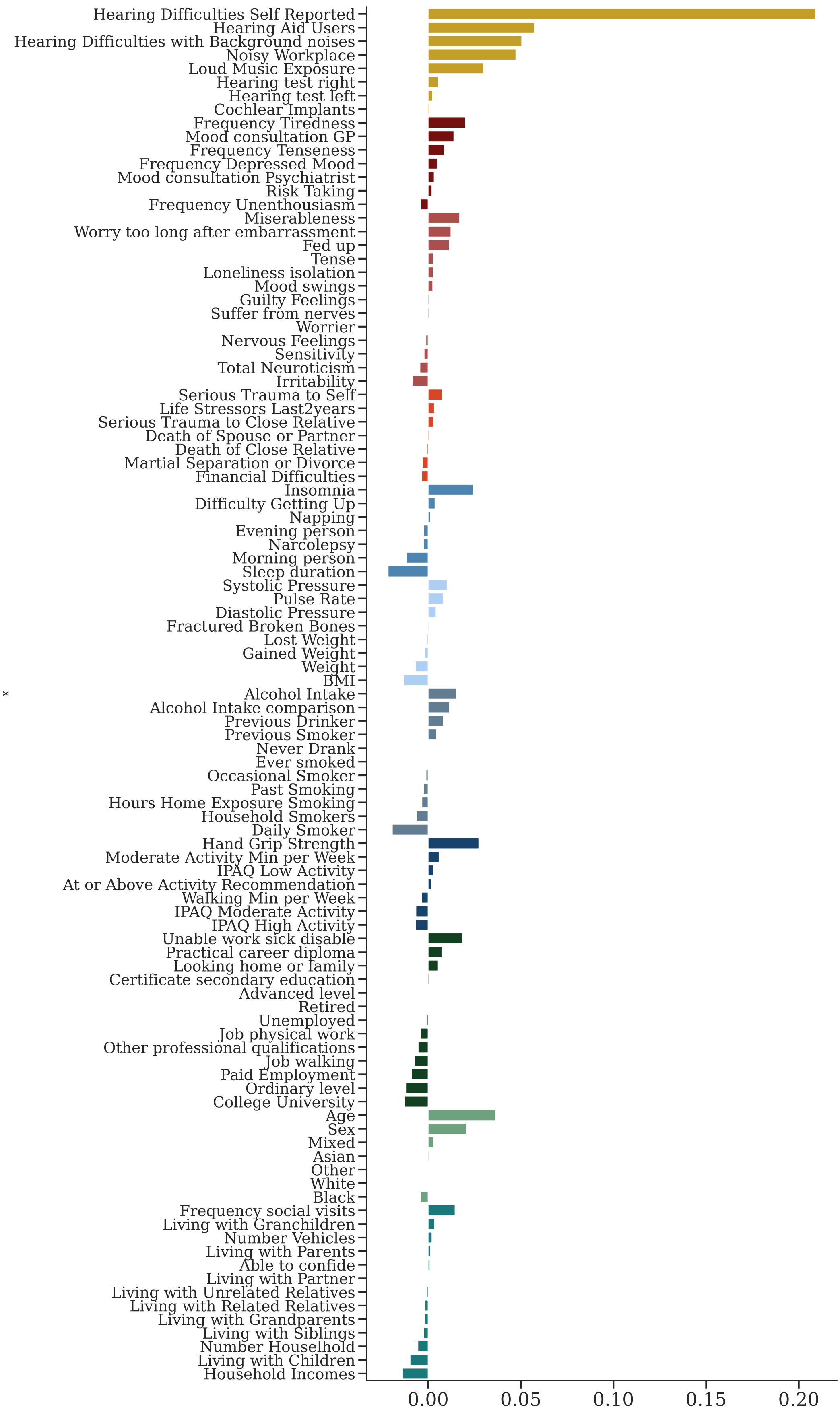

**Figure A2:** Features loads (Pearson's *r* correlation coefficient between features and the models' scores) of the Presence Risk Score model

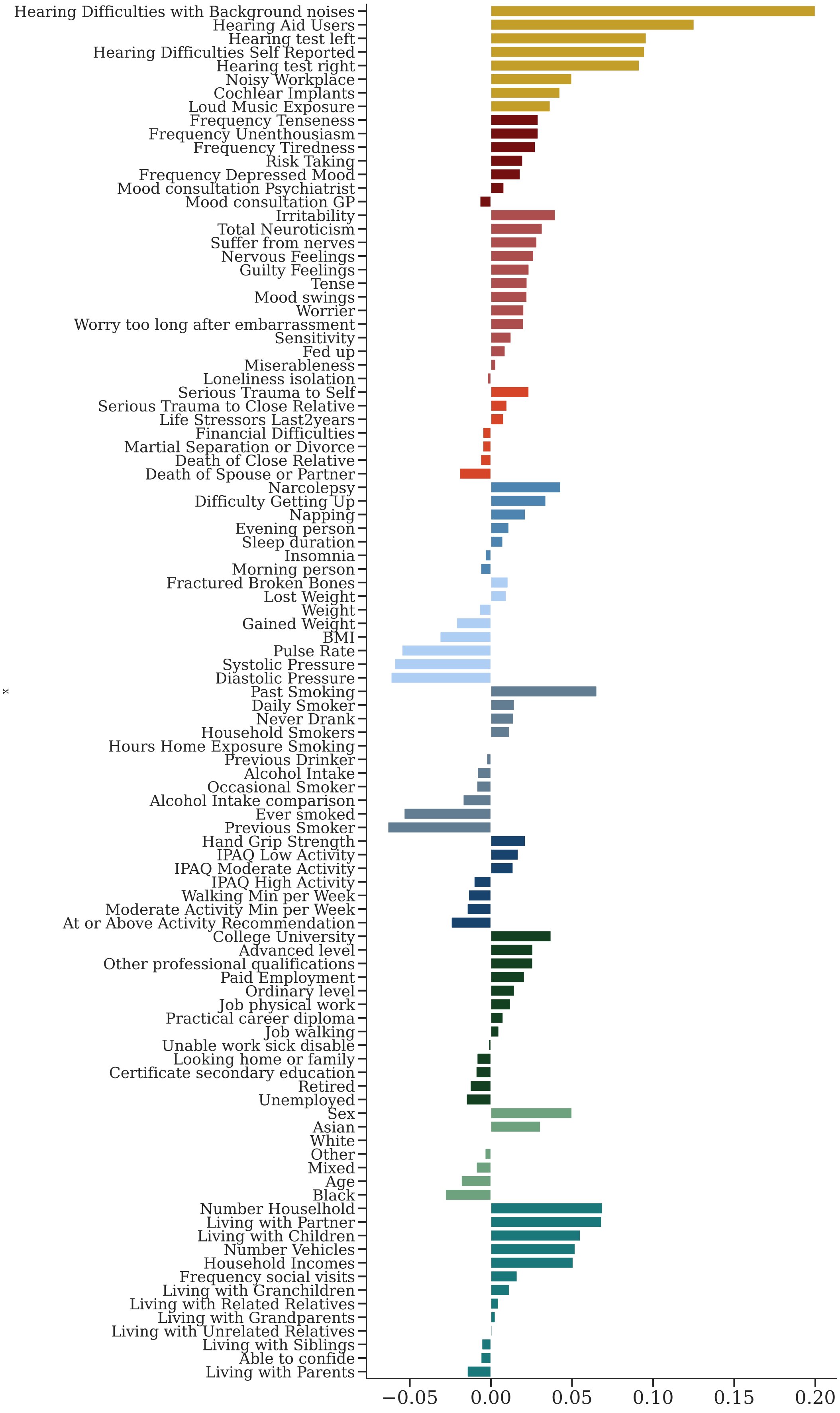

**Figure A3:** Features weights of the Severity Risk Score model

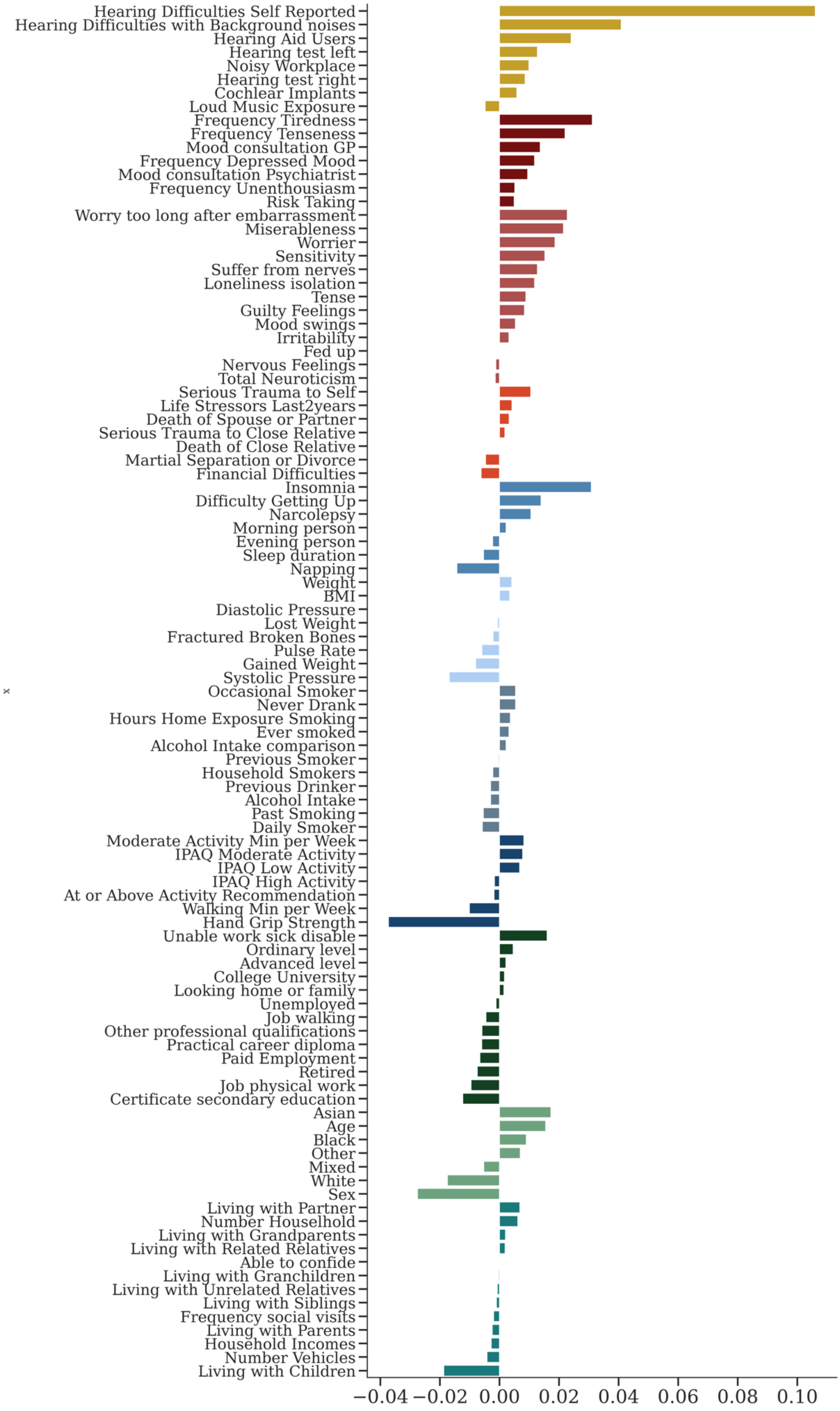

**Figure A4:** Features loads (Pearson's *r* correlation coefficient between features and the models' scores) of the Severity Risk Score model

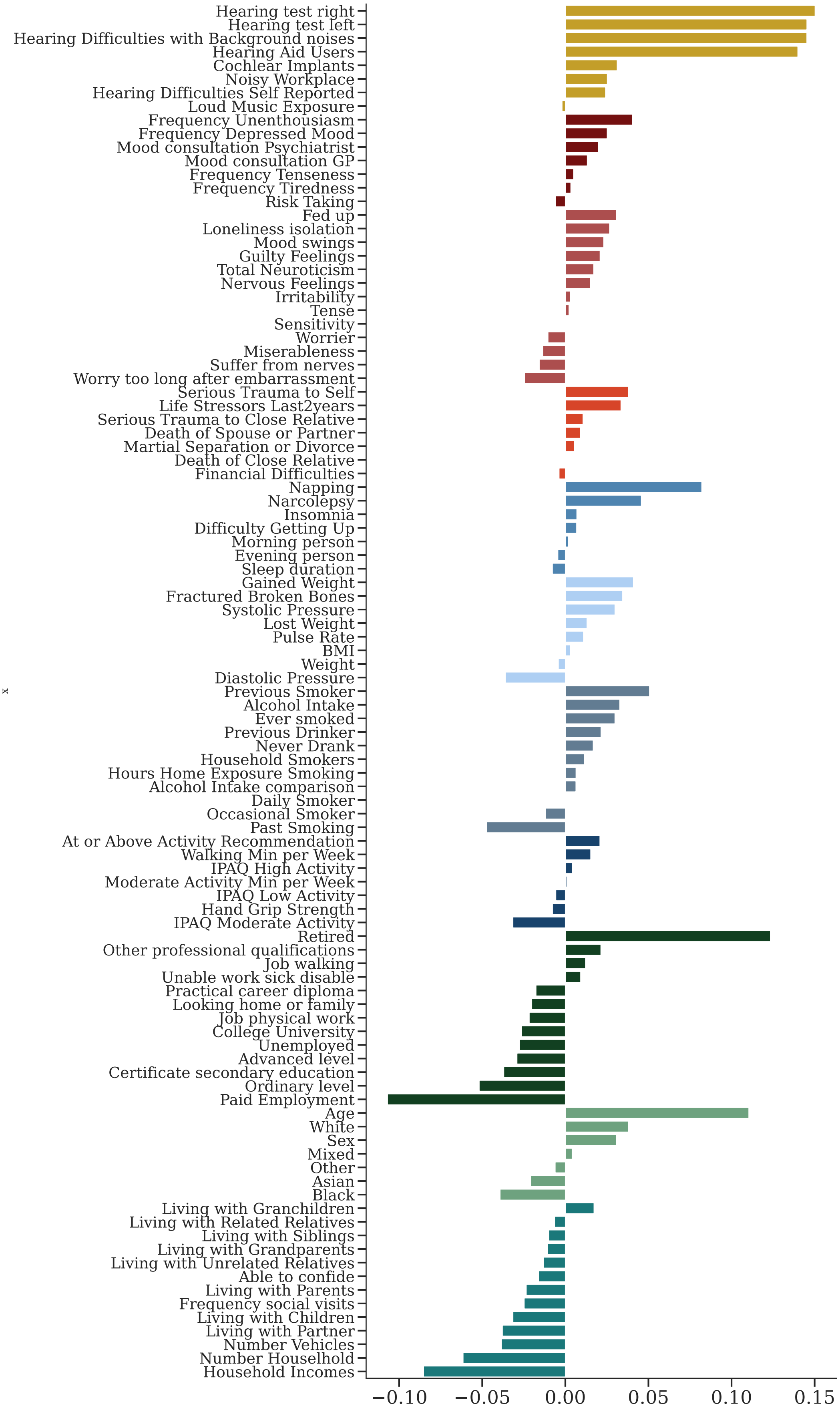
